# A Clinical Study to Assess the Safety and Pharmacokinetics of Orally Administered Strontium *L*-lactate in Healthy Adults

**DOI:** 10.1101/2025.07.15.25331565

**Authors:** Deanna J. Nelson, Kristen Sanoshy, Eunice Mah, the Biofortis Innovation Services Team

## Abstract

**Purpose:** Strontium salts may provide support for bone health and treatment for osteoporosis, but the safety and effectiveness of these salts is not completely understood. The aim of this clinical study (NCT03761979) was to obtain safety and pharmacokinetic information following acute oral intakes of three ascending doses of strontium *L*-lactate by healthy adults.

**Subjects Methods:** Ten healthy men and women, mean age 43 ± 2 years, ingested one of three ascending doses of strontium *L*-lactate (SrLac) once per week for three weeks in succession. All subjects were administered the Study Product in a sequential manner such that the lowest amount (170 mg Sr) was provided at Visit 2, the next highest amount (340 mg Sr) was provided at Visit 3, and the highest amount (680 mg Sr) was provided at Visit 4. At each visit, fasting blood collections were performed pre-dose and 1, 2, 3, 4, 5, 6, 8 and 12 hours post-dose to determine serum strontium at each interval.

**Results:** The pharmacokinetics related to each of three doses of SrLac that were administered to fasted subjects were similar to the findings for other strontium salts. At a dose of 170 mg strontium, a mean serum C_max_ of 2.6 ± 0.6 mg Sr/dL was observed about 3.1 h after ingestion. A dose of 340 mg strontium exhibited a mean serum C_max_ of 6.4 ± 1.8 mg Sr/dL about 3.2 h after ingestion. At a dose of 680 mg strontium, a mean serum C_max_ of 9.3 ± 2.1 mg Sr/dL was observed about 2.8 h after ingestion. Oral bioavailability was high, reflecting the high solubility of SrLac in water and intestinal fluid. The data suggest that between 27% and 34% of the administered dose was absorbed. At these doses, no strontium-related adverse effects were observed.

**Conclusions:** This clinical study in 10 normal adults (50% females) showed that the strontium ion in SrLac is readily bioavailable after oral administration. The intervention was conducted per study protocol, and no clinically significant protocol deviations occurred. Pharmacokinetic data indicated that doses of 170 and 340 mg strontium provided serum strontium concentrations in ranges known to be beneficial for the treatment of low bone density of osteoporosis and osteopenia. No product-related adverse events were observed.

## Introduction

Osteoporosis is a global health concern that continues to gain importance in the field of public health as the world’s population ages [1,2]. Increased bone fragility, which is the hallmark of osteoporosis, results from several factors including age-related hormonal changes in both men and women, use of glucocorticoids for alleviation of pain and treatment of autoimmune disorders, and changes to a more sedentary lifestyle with age [3,4]. Although recent estimates suggest that almost 53 million Americans, have osteoporosis or low bone density [5], many individuals are not aware of the disorder until an unexpected fracture of the wrist, hip or spine occurs. A fractured spine or hip is particularly devastating, since this break often leads to substantial losses in mobility, autonomy, and quality of life, in addition to increased mortality and huge societal costs. In fact, in the United States the direct cost of care resulting from hip or spine fractures is estimated to be between **$17 and 22 billion**. This estimate excludes indirect costs related to reductions in quality of life, productivity, and lifespan [6] showing that total costs related to this disease are likely to be significantly higher.

In general, bone health can be maintained throughout life [7,8] by adhering to certain dietary and lifestyle choices. Getting sufficient calcium and magnesium from the diet helps build and maintain strong bones. Vitamin D is also crucial to bone health because it upregulates calcium receptors and facilitates calcium utilization by the body. Thus, eating Vitamin D-rich foods and getting sufficient exposure to sunlight, which aids conversion of Vitamin D to its active form, are also important for maintaining bone health throughout life. Finally, regular weight-bearing exercises have also been shown to increase bone density.

If these actions are not sufficient to maintain bone health, several medicines are known to slow bone loss and treat osteoporosis when administered together with calcium and vitamin D. A strontium salt is one such medication [9-14]. Clinical studies in adults with osteoporosis have demonstrated that the strontium salt, strontium ranelate, reduces the incidence of fractures (both vertebral and non-vertebral) and increases bone mass and bone mineral density [15-19]. Other strontium salts (e.g., strontium *D,L*-lactate, strontium gluconate, strontium citrate, and strontium malonate) have been safely administered as supplements to both normal subjects and subjects with osteoporosis at a daily oral dose as high as 1.7 g strontium (Sr) and for dosing periods from several months to over a decade [9, 20-22]. Current data indicate that organic strontium salts such as these provide better support for bone health than inorganic salts [14]. Equally importantly, daily administration of a strontium salt to over 200,000 women for periods exceeding a decade has been shown to have low risk of potential side effects and has proven free of unexpected adverse events [22]. Thus, both historical and current data indicate that strontium salts have exciting potential as a means for preventing and treating the bone deterioration of osteopenia and osteoporosis.

Although strontium *D,L*-lactate was marketed as Strontolac® by Wyeth during the 1950’s, only one clinical report summarizes its benefits in treatment of osteoporosis [9]. Unlike strontium ranelate, a strontium-containing drug that has been extensively studied, no recent clinical trials have been conducted with strontium *D,L*-lactate or the strontium salt of the single *L*-enantiomer of lactic acid, SrLac. We selected SrLac because this salt is highly water-soluble source of strontium ion (Sr^2+^) and has no taste or odor, either as the solid or in solution. *L*-lactate is a natural, non-toxic anion. It was our position that previous findings justified further investigation into the therapeutic efficacy of SrLac against osteoporosis. Therefore, this clinical study was conducted to obtain general safety and pharmacokinetic (PK) information following acute oral intakes of three doses of SrLac by healthy adults [23].

## Material and Methods

### Study Product

The Study Product was a highly pure form of SrLac, the strontium salt of *L*-lactic acid. SrLac was manufactured by BioLink Life Sciences in compliance with current Good Manufacturing Practices. SrLac was thoroughly tested and met rigorous internal purity specifications. It was free from contamination by *D*-lactic acid and trace metals known to harm human health.

### Ethics

This study was conducted according to Good Clinical Practice (GCP) Guidelines (Title 21 of the U.S. Code of Federal Regulations) and the Declaration of Helsinki (2002). Signed written informed consent for participation in the study was obtained from all subjects before protocol-specific procedures were carried out.

Subjects were also informed of their right to withdraw from the study at any time. The study was explained verbally, as well as on the Institutional Review Board (IRB)-approved informed consent document. Each subject was given ample opportunity to inquire about details of the study and to read and understand the consent form before signing it. Consent was documented by the dated signature of the subject; each subject received a copy of the written informed consent document after signature. The informed consent document also included HIPAA-compliant wording, by which subjects authorized the use and disclosure of their Protected Health Information by the Investigator and by those persons who need that information for the purposes of this study. Subjects were also advised that the Sponsor, its employees or agents, the IRB, as well as representatives of the Food and Drug Administration (FDA), would have the right to audit and review pertinent medical records relating to this clinical trial as part of the informed consent.

Subjects’ anonymity was maintained on electronic case report forms (eCRFs) and other documents by utilization of initials, number, or code, and not by using a subject’s name. The Investigator kept a separate log showing codes, names, and addresses. All documents showing the subjects’ identity were kept in strict confidence by the Biofortis team.

The study protocol was approved by an Institutional Review Board (IntegReview, Austin, TX) prior to study commencement and subject recruitment.

### Subjects

Subjects (N = 10) were men (N = 5) and women (N = 5), 18 to 65 years of age, with BMIs that ranged from normal to obese (18.0 to 31.9 kg/m^2^). Each subject was assessed for eligibility according to the inclusion and exclusion criteria described in the Study Protocol [23 and Supplementary Material].

### Administration of Study Product

The Sponsor (BioLink Life Sciences) provided SrLac as a dry powder packaged in individual vials. The powder was prepared for consumption by adding 10 mL of distilled water to the vial and stirring until dissolution was complete. This liquid was then poured into an administration cup. An additional 10 mL of distilled water was used to rinse the remaining SrLac into the administration cup. Then 80 mL of distilled water was added directly into the administration cup. Following consumption of the 100 mL solution of SrLac, another 100 mL of distilled water was added into the administration cup, swirled, and consumed by the subject.

All subjects were administered the Study Product in a sequential manner such that the lowest amount (170 mg Sr) was provided at Visit 2, the next highest amount (340 mg Sr) was provided at Visit 3, and the highest amount (680 mg Sr) was provided at Visit 4.

### Study Design

This was an unblinded, sequential, three dose study (Figure 1). The study consisted of one screening visit (Visit 1; day -7) and three test visits (Visits 2, 3, and 4; days 0, 7, and 14) with at least a 6-d washout between test visits.

**Figure 1.**
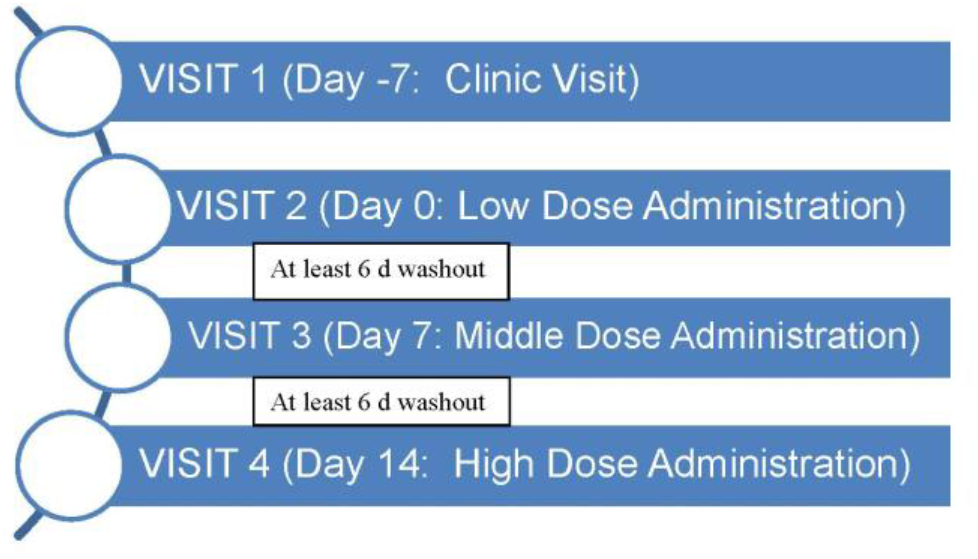
Study design.

At Visit 1 (day -7), subjects provided informed consent and underwent screening assessments including evaluations of medical history, prior/current medication/ supplement use, inclusion and exclusion criteria, height, body weight, body mass index (BMI), vital signs, and Vein Access Scale. A fasting capillary blood glucose was assessed to establish each subject’s fasting level and an in-clinic urine pregnancy test was also obtained for all women < 60 years of age. Additionally, a fasting blood sample was collected for a chemistry profile and hematology panel. Study instructions were also provided and included fasting compliance (water only) and avoidance of alcohol 3 d prior to test visits (Visits 2, 3, and 4; days 0, 7, and 14]. Subjects were dispensed a 24-h Diet Record with instructions to record intake the day prior to Visit 2 (day 0).

At Visit 2 (day 0), subjects arrived at the clinic fasted for the 12-h PK test visit. Subjects went through clinic visit procedures including: a review of inclusion/exclusion criteria, concomitant medication/supplement use, body weight, and vital signs assessment. Adverse events (AE) were assessed, and subjects were queried about compliance to study instructions. Additionally, a 24-h diet record was collected and reviewed. Fasting capillary blood glucose was assessed to confirm fasting status. Eligible subjects were enrolled and began the PK test with the insertion of an intravenous catheter at least 5 min prior to the first blood sampling time. In order to maintain patency of the intravenous catheter, the catheter was flushed with normal saline solution at least hourly. (Blood samples were drawn by venipuncture if the catheter failed.) Blood samples were obtained at t = -0.25 h ± 10 min for the PK analysis of serum strontium, where t = 0 h was the time of SrLac consumption. Additional blood samples were collected at all timepoints for backup. Subjects consumed the study product in its entirety within 10 min at t = 0 h. No food or water was ingested for the subsequent 2-h period. Blood samples were obtained for the PK analysis of serum strontium at t = 1 and 2 h ± 5 min, where t = 0 h was the start of study product consumption. Subjects were administered a standard breakfast immediately following the t = 2 h blood draw. *Ad libitum* water consumption was allowed for the remainder of the visit following study product consumption. Water consumption was recorded for replication at the subsequent test visits. Blood samples for serum strontium analysis were obtained via the indwelling venous catheter or venipuncture at t = 3, 4, 5, 6, and 8 h ± 5 min, where t = 0 h was the start of study product consumption. Subjects were provided lunch (immediately following the t = 6 h blood draw), a snack (immediately following the t = 8 h blood draw) and dinner (at t = 10 h). Following the final blood draw at t = 12 h ± 5 min, adverse events (AEs) were assessed. Subjects were dispensed a blank/new 24-h Diet Record to record all food and beverage consumed the day prior to Visit 3 (day 7); as well as a copy of the completed 24-h Diet Record (from day -1) with instructions to replicate the same food and beverage intake as closely as possible the day prior to the subsequent visit. Duplicate study instructions were also provided.

At Visits 3 and 4 (days 7 and 14), subjects returned to the clinic for clinic visit procedures (i.e., vital signs; body weight; review inclusion and exclusion criteria for relevant changes, and concomitant medication/supplement use review). AEs were assessed and subjects were queried about study instructions compliance. The 24-h Diet Record was collected, and study staff reviewed the 24-h Diet Record to compare food and beverage consumption to the day -1 recall for consistency. If the subject’s food intake was not consistent from the 24-h Diet Record reviewed at Visit 2 (day 0), the study staff contacted the Project Manager before continuing the visit as the test visit may have required rescheduling. Subjects then continued to the next sequential dose of SrLac [medium dose at Visit 3 (day 7) and high dose at Visit 4 (day 14)] and repeated the PK test described above for Visit 2.

Compliance was defined as consuming each dose of SrLac in its entirety during the test visit, per protocol.

### Primary Outcome Variable

The primary outcome variable was the incremental area under the curve (iAUC) for serum strontium from pre-product consumption (t = -0.25 h) to 12 h (iAUC_-0.25-12h_). See Statistical Analyses for further details.

### Secondary Outcome Variables

Secondary outcome variables included the following parameters:

- iAUC for serum strontium from pre-product consumption (t = -0.25 h) to infinity (iAUC_-0.25-∞_)
- Maximum serum concentration (C_max_)
- Time to C_max_ (T_max_)
- Rate of elimination (K)
- Half-life (t_1/2_)
- Oral bioavailability (F) and the fraction of the amount of strontium given orally that reaches the systemic circulation

### Clinical and Laboratory Assessments

Measurements included the Clinic Visit Assessments, during which height (first visit only), BMI assessment (first visit only), vital signs, body weight, evaluation of inclusion/exclusion criteria and concomitant medication use were conducted. Standardized vital signs measurements included resting blood pressure and pulse measured using an automated blood pressure measurement device. Blood pressure was obtained after the subject had been sitting for at least five min. Systolic and diastolic pressures were measured once using an appropriately sized cuff. The Accu-Check^®^ Performa (Roche, Indianapolis, IN) finger-stick glucometer was used for determination of the in-clinic (Biofortis Clinical Research, Addison, IL) fasting capillary blood glucose at Visit 1, 2, 3, and 4 (days -7, 0, 7, and 14) to ensure adequate fasting (10-14 h).

At Visit 1 (day -7), the fasting (10-14 h) chemistry profile was measured, including: glucose, sodium, potassium, chloride, carbon dioxide, BUN, creatinine, calcium, osmolality, AST, ALT, ALP, total bilirubin, total protein, and albumin. In addition, the fasting (10-14 h) hematology measurements included: WBC, RBC, hemoglobin concentration, hematocrit (as volume percent), MCV, MCH, MCHC, neutrophils, lymphocytes, monocytes, eosinophils, basophils and platelet count. [Analytes were assessed by Elmhurst Memorial Reference Laboratory (Elmhurst, IL).] Lastly, an in-clinic urine pregnancy test was performed on all women <60 years of age during this visit.

Blood samples for the analysis of serum strontium were collected via intravenous catheter at Visits 2, 3 and 4 (days 0, 7, and 14) at t = -0.25 h (± 10 min) and t = 1, 2, 3, 4, 5, 6, 8, 10, and 12 h (± 5 min), where t = 0 h was the start of study product consumption. The serum strontium concentrations were determined by NMS Labs (Willow Grove, PA). Additional samples were stored as backup in the event the original sample was not viable.

Subjects recorded all food and beverages consumed the day prior to Visit 2 (day 0) and were instructed to replicate the same diet intake the day prior to Visits 3 and 4 (days 7 and 14).

### Safety Assessments

Safety was assessed by evaluation of AEs, as well as measurement of vital signs and body weight.

### Statistical Analyses

The statistical methods and data analyses were conducted by a Biofortis statistician using SAS for Windows® (version 9.2). Descriptive statistics (number of subjects, mean, standard error of the mean (SEM), median, inter-quartile limits, minimum, and maximum values) were presented for subject demographics and anthropometric measurements collected at screening/baseline. Descriptive statistics (i.e. number of subjects, minimum and maximum, median, interquartile limits, mean, and SEM) were presented for all the continuous outcomes for each dose level. The data were also used to prepare a time response graph depicting mean and SEM of serum strontium concentrations for each dose level. Incremental AUC was calculated for serum strontium, using the linear trapezoidal rule of the standard non-compartmental analysis [24]. Rate of elimination, half-life and iAUC_-0.25-∞_ were calculated as previously described [25].

## Results

Selected demographics and baseline characteristics for all enrolled subjects are summarized in **Table 1**.

**Table 1.** Characteristics of Study Participants.

| **Characteristic** |  | **Overall (N=10)** |
| --- | --- | --- |
| Gender | Male | 5 (50.0%) |
|  | Female | 5 (50.0%) |
| Age (years) | Mean (SEM) | 43.50 (2.91) |
|  | Standard Deviation | 9.22 |
|  | Median | 41.00 |
|  | Q1, Q3 | 37.00, 51.00 |
|  | Minimum, Maximum | 33.00, 61.00 |
| Ethnicity | Hispanic or Latino | 1 (10.0%) |
|  | Not Hispanic or Latino | 9 (90.0%) |
| Race | Asian | 1 (10.0%) |
|  | White | 9 (90.0%) |
| Weight (kg) | Mean (SEM) | 73.40 (2.98) |
|  | Standard Deviation | 9.41 |
|  | Median | 69.90 |
|  | Q1, Q3 | 66.10, 79.20 |
|  | Minimum, Maximum | 64.50, 91.10 |
| Height (cm) | Mean (SEM) | 170.61 |
|  | Standard Deviation | 7.24 |
|  | Median | 170.45 |
|  | Q1, Q3 | 165.80, |
|  | Minimum, Maximum | 161.50, |
| BMI (kg/m<sup>2</sup>) | Mean (SEM) | 25.17 (0.74) |
|  | Standard Deviation | 2.35 |
|  | Median | 25.00 |
|  | Q1, Q3 | 23.80, 26.10 |
|  | Minimum, Maximum | 21.30, 29.00 |

**Abbreviations:** N, sample size; SEM, standard error of the mean; Q1, quartile 1; Q3, quartile 3

### Pharmacokinetic Data

Summary results for the incremental area under the curve for serum strontium from pre-product consumption to 12 h (the primary outcome) for the sample population (N = 10) are shown in **Table 2**.

**Table 2.**
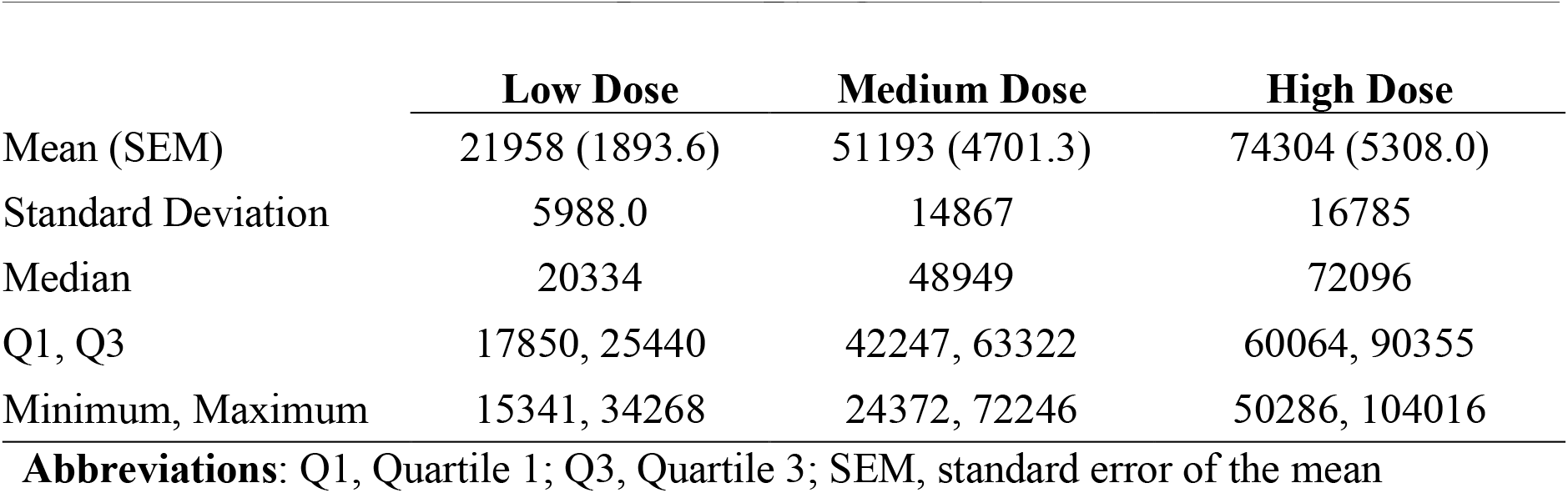
Serum Strontium iAUC-_0.25h - 12h_ (mcg/L × h)

|  | Low Dose | Medium Dose | High Dose |
| --- | --- | --- | --- |
| Mean (SEM) | 21958 (1893.6) | 51193 (4701.3) | 74304 (5308.0) |
| Standard Deviation | 5988.0 | 14867 | 16785 |
| Median | 20334 | 48949 | 72096 |
| Q1, Q3 | 17850, 25440 | 42247, 63322 | 60064, 90355 |
| Minimum, Maximum | 15341, 34268 | 24372, 72246 | 50286, 104016 |

**Abbreviations:** Q1, Quartile 1; Q3, Quartile 3; SEM, standard error of the mean

Summary results for the secondary outcome variables are shown in **Tables 3-7**. As expected, serum Sr concentrations at 12 hours, as well as the maximum observed serum concentrations, increased as the dosage increased (Tables 3 and 4). The time needed to reach the maximum strontium serum concentration and rates of strontium elimination were not significantly different between dosages (Tables 5 and 6). The half-life of Sr was approximately 2 hours longer in the high dosage compared to the low and medium doses (Table 7).

**Table 3.** Serum Strontium iAUC-_0.25h - infinity_ (mcg/L × h)

|  | Low Dose | Medium Dose | High Dose |
| --- | --- | --- | --- |
| Mean (SEM) | 47872 (4168.4) | 115632 (10765) | 185030 (13336) |
| Standard Deviation | 13182 | 34042 | 42173 |
| Median | 43534 | 122900 | 188848 |
| Q1, Q3 | 38332, 55116 | 84631, 147262 | 152477, 198916 |
| Minimum, Maximum | 34518, 77257 | 67011, 151475 | 125540, 266138 |

**Abbreviations:** Q1, Quartile 1; Q3, Quartile 3; SEM, standard error of the mean

**Table 4.** Maximum Serum Strontium Concentration (C_max_) (mcg/L)

|  | Low Dose | Medium Dose | High Dose |
| --- | --- | --- | --- |
| Mean (SEM) | 2620.0 (205.91) | 6380.0 (567.41) | 9320.0 (675.08) |
| Standard Deviation | 651.15 | 1794.3 | 2134.8 |
| Median | 2500.0 | 5950.0 | 9050.0 |
| Q1, Q3 | 2200.0, 3200.0 | 5200.0, 8100.0 | 7500.0, 11000 |
| Minimum, Maximum | 1800.0, 3800.0 | 3200.0, 8500.0 | 6600.0, 13000 |

**Abbreviations:** Q1, Quartile 1; Q3, Quartile 3; SEM, standard error of the mean

**Table 5.**
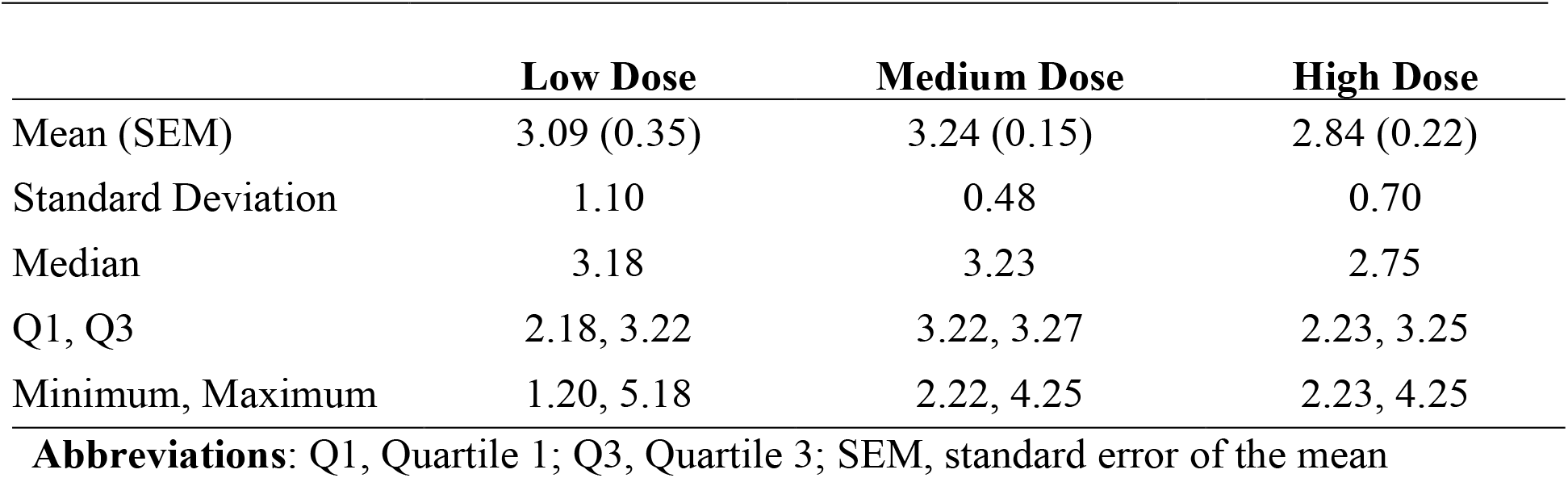
Serum Strontium Time to C_max_ (T_max_) (h)

**Table 6.** Serum Strontium Rate of Elimination (h^-1^)

|  | Low Dose | Medium Dose | High Dose |
| --- | --- | --- | --- |
| Mean (SEM) | 0.06 (0.00) | 0.06 (0.00) | 0.05 (0.00) |
| Standard Deviation | 0.01 | 0.01 | 0.01 |
| Median | 0.05 | 0.06 | 0.05 |
| Q1, Q3 | 0.05, 0.07 | 0.05, 0.07 | 0.04, 0.06 |
| Minimum, Maximum | 0.04, 0.08 | 0.04, 0.08 | 0.04, 0.08 |

**Abbreviations:** Q1, Quartile 1; Q3, Quartile 3; SEM, standard error of the mean

**Table 7.**
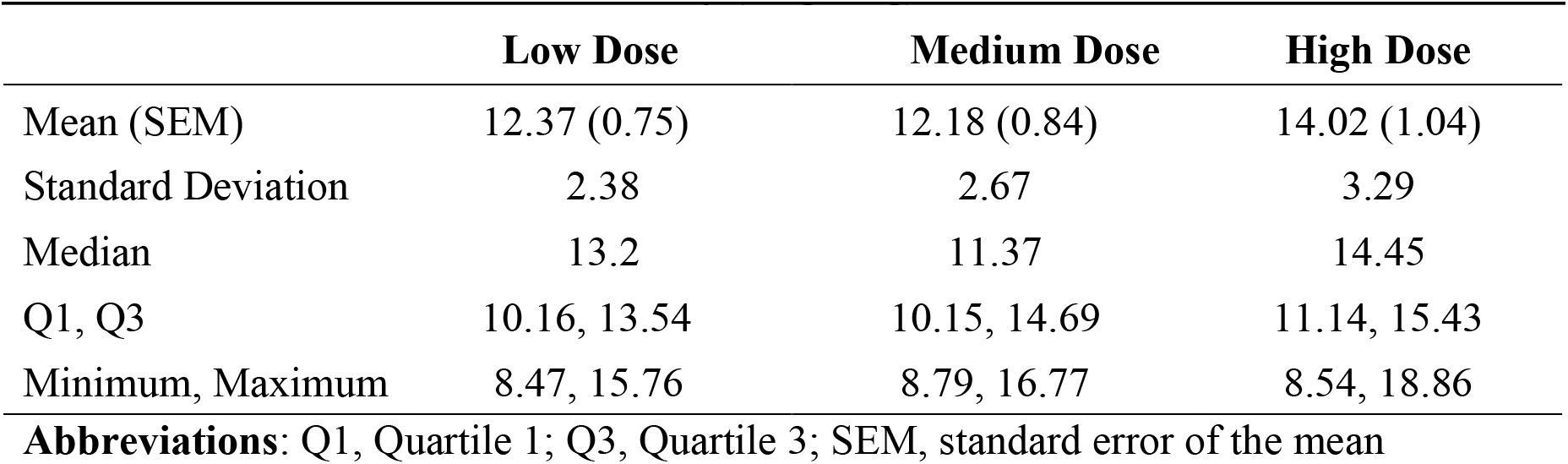
Serum Strontium Half life (t_1/2_) (h)

Summary results for strontium oral bioavailability for the sample population (N = 9-10) are shown in **Table 8**. Results from two different estimates are presented.

The absolute oral bioavailability of an orally administered product can only be obtained in the presence of PK data from intravenous administration of the product at the same dose. To our knowledge, no studies have calculated the volume of distribution (V) for strontium *L*-lactate in humans. Thus, in the absence of data from intravenous administration, oral bioavailability was calculated using the method of residuals, whereby V was estimated to be 64 L, consistent with that obtained from a study on another water-soluble strontium salt (i.e., strontium gluconate) [26]. Most of the oral bioavailability values calculated using this method were above 100, which would suggest that more than 100% of the strontium *L*-lactate consumed reached the systemic circulation. Since this is not mathematically possible it is likely that the V utilized to estimate F (i.e., V = 64 L) is too low.

Alternatively, oral bioavailability was calculated using a volume of distribution V calculated as the X_0_/intercept

where X_0_ = amount of study product provided orally (i.e. dose)

Intercept = y-intercept of the extrapolated line of the elimination curve

(The formula V = X_0_/intercept is commonly used for single intravenous bolus administration.)

Both calculations of oral bioavailability (either with V = 64 L or V = X_0_/intercept) required the calculation of the rate of absorption, Ka, as shown below:

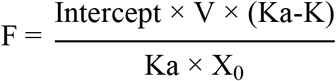

where F = oral bioavailability

Intercept = y-intercept of the extrapolated line of the elimination curve

V = volume of distribution Ka

= rate of absorption

K = rate of elimination

X_0_ = amount of study product provided orally (i.e. dose)

Ka was calculated using the method of residuals. The difference between the observed concentrations and the extrapolated concentration values prior to C_max_ (i.e., the residuals) was calculated and plotted on a semi-log graph to obtain the feathered or residual line. The slope of this line was then used to calculate Ka as follows: slope of residual line or absorption phase = e^(−Ka). Ka was successfully obtained for all subjects at all study product amounts except for one subject at 170 mg and 680 mg Sr. At these doses, the residuals were negative. Because it is not possible to take the log of negative numbers, the slope of the absorption phase cannot be calculated and thus, the rate of absorption cannot be obtained. These data were treated as missing values.

**Table 8.** Strontium Oral Bioavailability (mcg/mcg)

|  |  | Low Dose | Medium Dose | High Dose |
| --- | --- | --- | --- | --- |
| N |  | 9<sup>1</sup> | 10 | 9<sup>1</sup> |
| Oral bioavailability (F)<sup>2</sup> | Mean (SEM) | 1.08 (0.09) | 1.26 (0.12) | 0.92 (0.07) |
|  | Standard Deviation | 0.28 | 0.39 | 0.22 |
|  | Median | 1.06 | 1.19 | 0.93 |
|  | Q1, Q3 | 0.85, 1.23 | 1.07, 1.55 | 0.77, 1.05 |
|  | Minimum, Maximum | 0.70, 1.50 | 0.56, 1.84 | 0.65, 1.26 |
| Oral bioavailability (F)<sup>3</sup> | Mean (SEM) | 0.94 (0.01) | 0.93 (0.01) | 0.94 (0.01) |
|  | Standard Deviation | 0.04 | 0.03 | 0.02 |
|  | Median | 0.94 | 0.94 | 0.94 |
|  | Q1, Q3 | 0.93, 0.96 | 0.93, 0.95 | 0.94, 0.95 |
|  | Minimum, Maximum | 0.85, 0.97 | 0.86, 0.97 | 0.89, 0.96 |

**Abbreviations:** N, sample size; Q1, Quartile 1; Q3, Quartile 3; SEM, standard error of the mean
<sup>1</sup> Oral bioavailability for one subject at 170 mg Sr and 680 mg Sr doses cannot be obtained because the rate of absorption that is needed to calculate the ratio of volume of distribution to oral bioavailability cannot be calculated. These are treated as missing data.
<sup>2</sup> Oral bioavailability was calculated using volume of distribution (V) = 64 L.
<sup>3</sup> Oral bioavailability was calculated using volume of distribution (V) = $X_0/\text{intercept}$ , whereby $X_0$ is the amount of study product provided orally (i.e., dose) and *intercept* is the y-intercept of the extrapolated line of the elimination curve.

The fraction of the administered dose of SrLac that was absorbed was estimated as the ratio of iAUC_-0.25-∞_ to the administered dose (Table 9).

**Table 9.** Fraction of Administered Dose That was Absorbed.

|  | Low Dose, mg | Medium Dose, mg | High Dose, mg |
| --- | --- | --- | --- |
| Mean iAUC<sub>-0.25-∞</sub> | 47.872 | 115.632 | 185.030 |
| Administered Dose | 170 | 340 | 680 |
| Fraction Absorbed | 28% | 34% | 27% |

Fractional absorption values are comparable to the fractional absorption of other strontium salts [27–30].

### Strontium Metabolism

Overall time profile graphs of serum Sr concentration for each dose of study product are shown in Figure 2. The profiles are similar to those of other strontium salts [21,27-30]. (The strontium ion is not further metabolized. It binds to bone or is excreted in urine [10, 12-13].)

**Figure 2.**
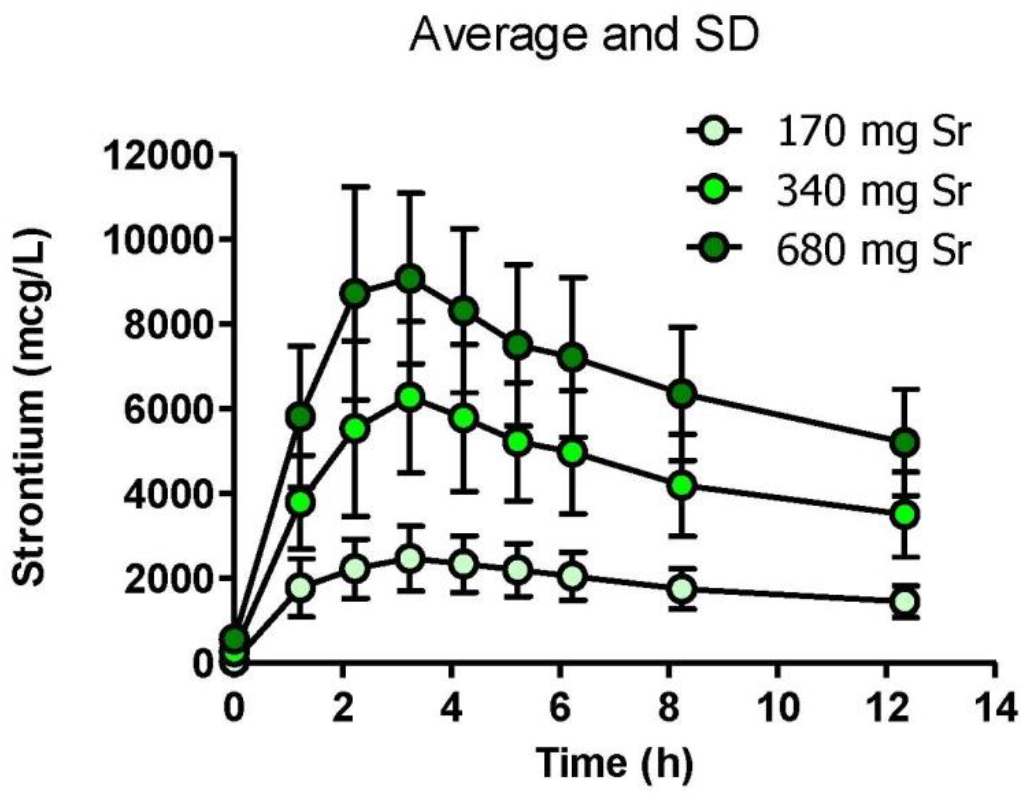
Time profiles of changes in serum strontium concentration.

### Safety

No clinical observation was judged to be related to study product. No AEs were observed.

## Discussion

In a 1950’s study at the Mayo Clinic, 72 patients with osteoporosis (aged 34-78 years) received over 6 g of strontium (DL-)lactate (1.7 g of Sr) daily with other medications for a period of 3 months to 3 years [9]. Of the 32 patients observed over the 3-year period, 27 (84%) showed significant improvement in mobility, including restoration in bone mineral density and freedom from pain. No adverse effects were reported, and no serious adverse effects were observed. However, given current knowledge, this sizeable daily dose of strontium appears unreasonable.

The data summarized in this report indicate that strontium *L*-lactate (SrLac) can be included in the group of strontium salts that provide support for bone health at significantly lower daily doses than were administered historically. In this study, 10 healthy men and women with a mean age 43, SD ± 2 years, ingested one of three ascending doses of SrLac once per week for 3 weeks in succession. Fasting blood collections were performed pre-dose and 1, 2, 3, 4, 5, 6, 8 and 12 hours post-dose to determine serum strontium at each interval.

The pharmacokinetics related to each of three doses of SrLac that were administered to fasted subjects were similar to the findings for other strontium salts. At a dose of 170 mg strontium, a mean serum C_max_ of 2.6 ± 0.6 mg Sr/dL was observed about 3.1 h after ingestion. A dose of 340 mg strontium exhibited a mean serum C_max_ of 6.4 ± 1.8 mg Sr/dL about 3.2 h after ingestion. At a dose of 680 mg strontium, a mean serum C_max_ of 9.3 ± 2.1 mg Sr/dL was observed about 2.8 h after ingestion. Pharmacokinetic data indicated that doses of 170 and 340 mg strontium provided serum strontium concentrations in ranges known to be beneficial for the treatment of low bone density of osteoporosis and osteopenia. Oral bioavailability was high, reflecting the high solubility of SrLac in water and intestinal fluid. Data suggest that between 27% and 34% of the administered dose was absorbed. At these doses, no strontium-related adverse effects were observed.

## Strengths and Limitations

Our study is the first to evaluate bioavailability of the water-soluble strontium salt strontium *L*-lactate in men and women. Although the size of the study cohort (10 adults) is small, the data mirror those provided in other reports of strontium bioavailability [27-30].

## Conclusions

For over a century, strontium (Sr) has been recognized as a “bone-seeking” ion [31-33]. In reflection of this activity, a number of strontium salts have been used as treatments for bone disorders, including osteopenia, osteoporosis, and bone pain. The strontium ion (Sr^2+^), the active ingredient in these salts, is absorbed paracellularly from the intestinal lumen [34]. The ion rapidly redistributes to bone. (Excess strontium is excreted in urine and feces.) Uptake of strontium by bone occurs preferentially in sites of active osteogenesis [35].

This clinical study in normal adults showed that the strontium ion in strontium *L*-lactate is bioavailable after oral administration. Pharmacokinetic data indicated that doses of 170 and 340 mg strontium provided serum strontium concentrations in ranges known to be beneficial for the treatment of low bone mineral density. No product-related adverse events were observed. Thus, strontium *L*-lactate may provide a viable option to support bone health throughout an individual’s lifetime.

## Data Availability

All data produced in the present study are available upon reasonable request to the authors.

## Acknowledgement

The authors wish to acknowledge the Biofortis Team. The abstract of this paper was presented at the 2018 Interdisciplinary Osteoporosis Symposium (New Orleans, LA) as a poster presentation. The full paper has never been published.

## References

1. Bongaarts J. Global fertility and population trends. Semin Reprod Med. 2015 Jan; 33(1): 5–10. doi: 10.1055/s-0034-1395272. PubMed PMID: 25565505.

2. Office of the Surgeon General (US). Chapter 4. The frequency of bone disease. In: Bone Health and Osteoporosis: A Report of the Surgeon General. Rockville (MD): Office of the Surgeon General (US); 2004. Available at: https://www.ncbi.nlm.nih.gov/books/NBK45515/.

3. Xiao PL, Cui AY, Hsu CJ, Peng R, Jiang N, Xu XH, Ma YG, Liu D, Lu HD. Global, regional prevalence, and risk factors of osteoporosis according to the World Health Organization diagnostic criteria: a systematic review and meta-analysis. Osteoporos Int. 2022 Oct;33(10):2137–2153. doi: 10.1007/s00198-022-06454-3. PMID: 35687123.

4. Weinstein RS. Glucocorticoids, osteocytes, and skeletal fragility: The role of bone vascularity. Bone 2010 Mar; 46(3): 564-70. PubMed PMID: 19591965; PubMed Central PMCID: PMC2823999.

5. GBD 2021 Diseases and Injuries Collaborators. Global incidence, prevalence, years lived with disability (YLDs), disability-adjusted life-years (DALYs), and healthy life expectancy (HALE) for 371 diseases and injuries in 204 countries and territories and 811 subnational locations, 1990-2021: a systematic analysis for the Global Burden of Disease Study 2021. Lancet. 2024 May 18;403(10440):2133–2161. doi: 10.1016/S0140-6736(24)00757-8. PMID: 38642570; PMCID: PMC11122111.

6. GBD 2021 Diseases and Injuries Collaborators. Global incidence, prevalence, years lived with disability (YLDs), disability-adjusted life-years (DALYs), and healthy life expectancy (HALE) for 371 diseases and injuries in 204 countries and territories and 811 subnational locations, 1990-2021: a systematic analysis for the Global Burden of Disease Study 2021. Lancet. 2024 May 18;403(10440):2133–2161. doi: 10.1016/S0140-6736(24)00757-8. Epub 2024 Apr 17. PMID: 38642570; PMCID: PMC11122111.

7. Levinger I, Brennan-Speranza TC, Zulli A, Parker L, Lin X, Lewis JR, Yeap BB. Multifaceted interaction of bone, muscle, lifestyle interventions and metabolic and cardiovascular disease: role of osteocalcin. Osteoporos Int 2017 Aug; 28(8): 2265-2273. PubMed PMID: 28289780.

8. Hart NH, Newton RU, Tan J, Rantalainen T, Chivers P, Siafarikas A, Nimphius S. Biological basis of bone strength: anatomy, physiology and measurement. J Musculoskelet Neuronal Interact. 2020 Sep 1;20(3):347-371. PMID: 32877972; PMCID: PMC7493450.

9. McCaslin FE, Janes JM. The effect of strontium lactate in the treatment of osteoporosis. Proc Staff Mtgs Mayo Clinic 1959; 34(13): 329–334.

10. Pilmane M, Salma-Ancane K, Loca D, Locs J, Berzina-Cimdina L. Strontium and strontium ranelate: Historical review of some of their functions. Mater Sci Eng C Mater Biol Appl. 2017 Sep 1; 78: 1222-1230. PubMed PMID: 28575961.

11. Maria S, Swanson MH, Enderby LT, D’Amico F, Enderby B, Samsonraj RM, Dudakovic A, van Wijnen AJ, Witt-Enderby PA. Melatonin-micronutrients Osteopenia Treatment Study (MOTS): A translational study assessing melatonin, strontium (citrate), vitamin D3 and vitamin K2 (MK7) on bone density, bone marker turnover and health related quality of life in postmenopausal osteopenic women following a one-year double-blind RCT and on osteoblast-osteoclast co-cultures. Aging 2017; 9(1): 256–285.

12. Marx D, Rahimnejad Yazdi A, Papini M, Towler M. A review of the latest insights into the mechanism of action of strontium in bone. Bone Rep. 2020 Apr 24;12:100273. doi: 10.1016/j.bonr.2020.100273. PMID: 32395571; PMCID: PMC7210412.

13. Kołodziejska B, Stępień N, Kolmas J. The Influence of Strontium on Bone Tissue Metabolism and Its Application in Osteoporosis Treatment. Int J Mol Sci. 2021 Jun 18;22(12):6564. doi: 10.3390/ijms22126564. PMID: 34207344; PMCID: PMC8235140.

14. Tomczyk-Warunek A, Turżańska K, Posturzyńska A, Kowal F, Blicharski T, Pano IT, Winiarska-Mieczan A, Nikodem A, Dresler S, Sowa I, Wójciak M, Dobrowolski P. Influence of Various Strontium Formulations (Ranelate, Citrate, and Chloride) on Bone Mineral Density, Morphology, and Microarchitecture: A Comparative Study in an Ovariectomized Female Mouse Model of Osteoporosis. Int J Mol Sci. 2024 Apr 6;25(7):4075. doi: 10.3390/ijms25074075. PMID: 38612883; PMCID: PMC11012416.

15. Cianferotti L, D’Asta F, Brandi ML. A review on strontium ranelate long-term antifracture efficacy in the treatment of postmenopausal osteoporosis. Ther Adv Musculoskelet Dis. 2013 Jun; 5(3): 127-39. PubMed PMID: 23858336; PubMed Central PMCID: PMC3707343.

16. Saidak Z, Marie PJ. Strontium signaling: molecular mechanisms and therapeutic implications in osteoporosis. Pharmacol Ther. 2012 Nov; 136(2): 216-26. PubMed PMID: 22820094.

17. Zacchetti G, Dayer R, Rizzoli R, Ammann P. Systemic treatment with strontium ranelate accelerates the filling of a bone defect and improves the material level properties of the healing bone. BioMed Res Int 2014; Article ID 549785.

18. Reginster JYl, Kaufman JM, Goemaere S, Devogelaer JP, Benhamou CL, FeIsenberg D, Diaz-Curiel M, Brandi ML, Badurski J, Wark J, Balogh A, Bruyere 0, Roux C. Maintenance of antifracture efficacy over 10 years with strontium ranelate in postmenopausal osteoporosis. Osteoporos Int. 2012 Mar; 23(3):1115–1122.

19. Khalid S, Calderon-Larrañaga S, Hawley S, Ali MS, Judge A, Arden N, van Staa T, Cooper C, Javaid MK, Prieto-Alhambra D. Comparative anti-fracture effectiveness of different oral anti-osteoporosis therapies based on “real-world” data: a meta-analysis of propensity-matched cohort findings from the UK Clinical Practice Research Database and the Catalan SIDIAP Database. Clin Epidemiol. 2018 Oct 9;10:1417–1431. doi: 10.2147/CLEP.S164112. PMID: 30349390; PMCID: PMC6183551.

20. Skoryna SC. Effects of oral supplementation with stable strontium. Canadian Med Assn J 1981; 125: 703–712.

21. Hansen C, Nilsson H, Christgau S, Andersen JEF. Water-soluble strontium salts for use in treatment of cartilage and bone conditions. U.S. Patent No. 7,595,342. Filed Nov 7, 2005. Date of patent Sept 29, 2009.

22. (a) Martín-Merino E, Petersen I, Hawley S, Álvarez-Gutierrez A, Khalid S, Llorente-Garcia A, Delmestri A, Javaid MK, Van Staa TP, Judge A, Cooper C, Prieto-Alhambra D. Risk of venous thromboembolism among users of different anti-osteoporosis drugs: a population-based cohort analysis including over 200,000 participants from Spain and the UK. Osteoporos Int. 2018 Feb;29(2):467–478. doi: 10.1007/s00198-017-4308-5. PMID: 29199359. (b) Ali MS, Berencsi K, Marinier K, Deltour N, Perez-Guthann S, Pedersen L, Rijnbeek P, Lapi F, Simonetti M, Reyes C, Van der Lei J, Sturkenboom M, Prieto-Alhambra D. Comparative cardiovascular safety of strontium ranelate and bisphosphonates: a multi-database study in 5 EU countries by the EU-ADR Alliance. Osteoporos Int. 2020 Dec;31(12):2425–2438. doi: 10.1007/s00198-020-05580-0. PMID: 32757044.

23. The Study Protocol has been published at ClinicalTrials.gov (NCT03761979).

24. Shiang, K-D. The SAS Calculations of Areas Under the Curve (AUC) for Multiple Metabolic Readings. SAS Conference Proceedings Western Users of SAS (WUSS). 2004. Available at: http://www.lexjansen.com/wuss/2004/posters/c_post_the_sas_calculations_.pdf

25. Matos-Pita AS, Bernardo de Miguel L. Non-compartmental pharmacokinetics and bioequivalence analysis. In: PharmaSUG 2005. (May 22-25, 2005, Phoenix, AZ). SAS Institute Inc., Pharmaceutical Industry SAS® Users Group (PharmaSUG). (Paper SP07). Available at: http://www.lexjansen.com/pharmasug/2005/statisticspharmacokinetics/sp07.pdf.

26. Moraes ME, Aronson JK, Grahame-Smith DG. Intravenous strontium gluconate as a kinetic marker for calcium in healthy volunteers. Br J Clin Pharmacol. 1991; 31(4):423–7.

27. Höllriegl V, Li WB, Oeh U, Roth P. Methods for assessing gastrointestinal absorption of strontium in humans by stable tracer techniques. Health Phys. 2006 Mar; 90(3):232–40. PubMed PMID: 16505620.

28. Höllriegl V, Louvat P, Werner E, Roth P, Schramel P, Wendler I, Felgenhauer N, Zilker T. Studies of strontium biokinetics in humans. Part 2: Uptake of strontium from aqueous solutions and labelled foodstuffs. Radiat Environ Biophys. 2002 Dec; 41(4): 281-7. Epub 2002 Dec 19.

29. Sips AJ, van der Vijgh WJ, Barto R, Netelenbos JC. Intestinal absorption of strontium chloride in healthy volunteers: pharmacokinetics and reproducibility. Br J Clin Pharmacol. 1996 Jun;41(6):543-9. PubMed PMID: 8799520; PubMed Central PMCID: PMC2042623.

30. Sips AJ, van der Vijgh WJ, Barto R, Netelenbos JC. Intestinal strontium absorption: from bioavailability to validation of a simple test representative for intestinal calcium absorption. Clin Chem. 1995 Oct;41(10):1446-50. PubMed PMID: 7586515.

31. Monograph: Strontium lactate. Merck’s 1907 Index. New York: Merck & Co., 1907. Page 425.

32. Papillon MF. Recherches experimentales sur les modifications de la composition immediate des os. C R Acad Sci 1870; 71: 372.

33. Skoryna SC. 1981. Effects of oral supplementation with stable strontium. Can Med Assoc J 125: 703–712.

34. Cabrera WE, Schrooten I, De Broe ME, D’Haese PC. Strontium and bone. J Bone Miner Res. 1999 May;14(5):661–8. doi: 10.1359/jbmr.1999.14.5.661. PMID: 10320513.

35. Querido W, Rossi AL, Farina M. The effects of strontium on bone mineral: A review on current knowledge and microanalytical approaches. Micron. 2016 Jan;80:122–34. doi: 10.1016/j.micron.2015.10.006. PMID: 26546967.

